# Prevalence of risky sexual behaviour and its association with alcohol use among unmarried adolescents and young adults in Eswatini: Evidence from a population-based survey

**DOI:** 10.1101/2025.11.24.25340940

**Authors:** Samkeliso G. Shongwe, Alexander W. Kay, Mduduzi C. Shongwe, Christopher O. Aimakhu, Thabile P. Simelane

## Abstract

HIV remains a major public health concern in sub-Saharan Africa (SSA), with adolescents and young adults (AYA) facing heightened vulnerability due to risky sexual behaviours (RSBs). In Eswatini, despite efforts to expand sexual and reproductive health (SRH) services, HIV prevalence and teenage pregnancy remain high. Alcohol use is a known driver of risky sexual behaviours, yet evidence on its association with risky sexual behaviour among AYA in Eswatini is limited. Therefore, this study aimed to examine the association between risky sexual behaviour and alcohol use among unmarried AYA in Eswatini. We conducted a cross-sectional analysis of the Swaziland HIV Incidence Measurement Survey 3 (SHIMS 3) data, using a weighted sample of 1,392 sexually active unmarried adolescents and young adults aged 15–24 years. We fitted logistic regression models to determine the association between risky sexual behaviour and alcohol use among unmarried adolescent and young adults in Eswatini with their 95% confidence interval (CI) at *p*-value < 0.05 statistical significance. The prevalence of risky sexual behaviour among AYA was 52.7% (95% CI: 50.1 – 55.3), while the prevalence of alcohol use was 31.3% (95% CI: 28.9 – 33.7). The study found a significant association between risky sexual behaviour and alcohol use (aOR = 2.35, 95% CI: 1.78 – 3.11). After controlling for covariates, residing in the Shiselweni region (aOR = 2.11, 95% CI: 1.55 – 2.86), and PrEP awareness (aOR = 1.60, 95% CI: 1.26 – 2.04) were associated with higher odds of engaging in risky sexual behaviour. This study found a significant association between risky sexual behaviour and alcohol use among unmarried adolescent and young adults in Eswatini. Addressing both substance use and sexual health through tailored, youth-friendly interventions is essential to reduce HIV transmission and improve SRH outcomes among adolescents and young adults.

## Introduction

Sub-Saharan Africa (SSA) continues to experience significant growth in the adolescents and young adults (AYA) population [1]. During adolescence and young adulthood, young people undergo major psychosocial, physiological, and cognitive changes that often increase sexual curiosity and the urge to explore and engage in new experiences [2]. These developmental changes often lead to increased engagement in risky sexual behaviours (RSBs) which contributes to negative sexual and reproductive health (SRH) outcomes, including unintended pregnancies, sexually transmitted infections (STIs), and HIV infection [3]. Risky sexual behaviour defined as any sexual activity that increases the chance of acquiring STIs and unwanted pregnancies. These activities include early sexual debut, having unprotected sex, having sex under the influence of alcohol, having multiple sexual partners, transactional sexual relationships and age-disparate sex [4]. Risky sexual behaviour among AYA is significantly more prevalent in developing countries, especially in SSA, than in developed countries [5]. These behaviours are a major driver in the spread of HIV, STIs, and unintended pregnancies among AYA in SSA, including Eswatini [6,7]. Unmarried AYA often face higher risks compared to their married counterparts, primarily due to barriers in accessing SRH services such as the absence of youth-friendly services, limited access to contraceptives, societal stigma, and poor sexual health education [8].

Eswatini continues to experience one of the highest HIV prevalence globally, with the epidemic posing a significant public health concern for adolescents and young adults aged 15–24 years. HIV prevalence is 5.6% among females aged 15–19 years, compared to 3.0% among males of the same age group. Among those aged 20–24 years, HIV prevalence is higher for females at 17.2%, compared to 3.9% for males [9]. STIs also continues to pose a serious public health concern among AYA in Eswatini [10]. Furthermore, teenage pregnancy remains another critical public challenge faced by young girls in Eswatini, with the adolescent birth rate at 87 births per 1,000 girls aged 15–19 years [11]. The continued spread of HIV, STIs, and teenage pregnancy among AYA in Eswatini is driven by a number of factors including inconsistence use of condoms, early initiation of sexual activities, involvement in substance use, and being involved with multiple sexual partners [7,12–15].

Alcohol consumption has been consistently identified as a major contributor to risky sexual behaviour among AYA [16], especially in high HIV burden settings like Eswatini. The country ranks as third-highest alcohol consumers globally, with a high per capita intake of alcohol [17], mostly practiced by young people. In Eswatini, heavy drinking among young people is often fueled by peer pressure, contributing to impaired judgment and lower inhibitions increasing vulnerability to sexual health risks such as unprotected sex, multiple sexual partnerships, sex with non-regular partners, and transactional sex [18,19], contributing to the burden of HIV, STIs and teenage pregnancy.

The government of Eswatini through the Ministry of Health in collaboration with various stakeholders have made significant progress in expanding access to HIV prevention and SRH services for young people, which includes implementation of school-based sexuality education, scaling up youth-friendly health services, promotion of voluntary medical male circumcision (VMMC), increasing uptake of pre-exposure prophylaxis (PrEP) and condom distribution [11,20–22]. Despite the progress made to strengthening the SRH services for young people in Eswatini, the country still reports high HIV prevalence, STIs and teenage pregnancy among young people, which reflects a pattern of unmet sexual and reproductive health needs.

Several studies have been conducted in Eswatini around risky sexual behaviours, however, these studies primarily focused on specific aspects, such as only adolescents aged 15-19 years, inconsistent condom use, timing of sexual debut, and intergenerational sexual partnerships [7,13,23,24]. There is limited evidence on the combined impact of various sexual risky behaviours (i.e., early sexual debut, condom use at last sex, sexual intercourse under the influence of alcohol and multiple sex partners) and its association with alcohol use among unmarried AYA in Eswatini. Therefore, the current study addressed this gap by estimating the prevalence of risky sexual behaviour and its association with alcohol use among AYA in Eswatini. Findings from this study are intended to inform, program designers, and stakeholders working on adolescent and youth health in Eswatini, aimed at improving HIV prevention, and access to youth-friendly SRH services thereby reducing the rates of new HIV infections, STIs and teenage pregnancy among AYA.

## Methods

### Study setting

The study was conducted in Eswatini, a small land locked country in the southern part of Africa. Eswatini has an estimated population of about 1.3 million people, characterized by a significant proportion of a youthful and growing population, with nearly half of the country’s population (46.9%) below the age of 25 [25]. Eswatini ranks among the countries with the highest HIV prevalence worldwide, with 24.8% of adults aged 15 and above living with HIV [9].

### Study design, data source and sampling procedures

This study employed a cross-sectional design using data from the 2021 Swaziland HIV Incidence Measurement Survey 3 (SHIMS 3), also known as the Eswatini Population-based HIV Impact Assessment 2021, conducted between May 2021 and November 2021. SHIMS 3 was a nationally representative, household-based survey targeting individuals aged 15 years and older, implemented as part of the multi-country Population-based HIV Impact Assessment (PHIA) initiative to measure the impact of Eswatini’s national HIV response. The SHIMS 3 survey collected data on the HIV epidemic status, HIV-related risk behaviours, coverage and impact of HIV services, and the country’s progress towards the UNAIDS 95-95-95 goals. SHIMS 3 used a two-stage, stratified cluster sampling design. The sampling frame included all households in Eswatini, based on the 2017 Population Census, covering 2,260 enumeration areas (EAs) and approximately 264,856 households. In the first stage, 200 EAs were selected stratified into 8 strata representing urban and rural areas across the 4 regions of Eswatini. Within each EA, households were randomly selected using equal probability sampling. On average, 35 households were chosen per cluster, with a range of 15 to 70 [9]. SHIMS 3 data is publicly accessible and can be requested online through the official PHIA data repository platform through https://phia-data.icap.columbia.edu/datasets. For this study the SHIMS 3 data was accessed from PHIA data repository platform on the 7^th^ of May 2025 and the data came already anonymized.

### Study population

The study population included all sexually active AYAs (aged 15 to 24 years) who were not married at the time of the survey. A weighted total of 1,392 AYAs were included in the analysis. AYAs who were married and sexually inactive were excluded from that analysis. AYAs with missing data on the variables of interest were also excluded from the analysis.

### Ethical consideration

This study used publicly available data from the PHIA data repository platform which comes already anonymized. Permission to get access to the data was obtained from the PHIA online request platform at: https://phia-data.icap.columbia.edu/datasets. PHIA surveys have been reviewed and approved by the U.S Centers for Disease Control and Prevention, Columbia University, Westat, as well as by the local ethics boards in the respective countries. This study also received approval and a waiver of informed consent from the Eswatini Health and Human Research Review Board (EHHRRB), Mbabane, Eswatini (Reference Number: EHHRRB 068/2025) because we were using secondary data. Participants provided consent during the time of the survey. The received dataset came anonymized to maintain its confidentiality.

## Measurement of variables

### Outcome variable

The outcome variable, risky sexual behaviour, was a composite measure derived from four indicators: (1) early sexual debut (*first sexual intercourse at age 14 or younger*); (2) unprotected sex during most recent sexual intercourse (*reporting no condom use during last sexual intercourse*); (3) having multiple sexual partners (*two or more sex partners in the past 12 months*); (4) alcohol use at last sexual encounter (*alcohol consumption by either the participant or their sexual partner during their most recent sexual encounter*). Respondents were classified as engaging in risky sexual behaviour if they reported one or more of these behaviours. The variable was coded as binary: ‘1’ for risky sexual behaviour, and ‘0’ for no risky sexual behaviour, as previously done in other studies [27].

### Main exposure variable

The main exposure variable for this study was alcohol use among participants. This was determined by asking participants to indicate the frequency with which they consume alcoholic beverages, with response options including: never, monthly or less, 2–4 times a month, 2–3 times a week, and 4 or more times a week. For the purpose of this study, participants who reported never consuming alcohol were classified as non-alcohol users, whereas those who selected any of the other categories indicating some level of alcohol consumption were classified as alcohol users, following the approach used in previous studies [28].

### Covariates

The study controlled for the following covariates: Age (15–19 years, and 20–24 years); sex (male, and female); educational status (primary or less, secondary/high school, and tertiary); place of residence (rural, and urban); region (Hhohho, Lubombo, Manzini, and Shiselweni); communicated with parent/guardian on SRH matters (yes, and no); contraceptive use (yes, and no); HIV testing (ever tested, and never tested); PrEP awareness (aware, and unaware); condom negotiation confidence (high confidence, moderate confidence, slight confidence, and no confidence).

## Operational definitions

### Communicated with parent/guardian on SRH matters

Communication with a parent or guardian on SRH issues was assessed based on two questions: whether AYA had ever talked with a parent or guardian about sex, and whether they had ever discussed HIV with a parent or guardian. AYAs who responded “Yes” to at least one of these questions were considered to have communicated with their parent or guardian on SRH matters. This classification is consistent with approaches used in previous studies [29].

### Contraceptive use

Contraceptive use was assessed based on the question of whether AYA or their sexual partners were currently doing something or using any method to delay or avoid pregnancy. AYAs who responded “Yes” were categorized as currently using contraception, while those who responded “No” were categorized as not using any contraceptive method.

### Awareness of pre-exposure prophylaxis (PrEP)

Awareness of pre-exposure prophylaxis (PrEP) was assessed by asking participants whether they had ever heard of PrEP prior to the survey. Those who responded “Yes” were categorized as being aware of PrEP, while those who responded “No” were considered unaware of PrEP.

### Condom negotiation confidence

Condom negotiation confidence was assessed by asking participants on their perceived ability and confidence to discuss and advocate for condom use with a sexual partner. Responses were categorized as: high confidence, moderate confidence, slight confidence, and no confidence, following the approach used in previous studies [30].

## Data management and statistical analysis

Data cleaning and analysis were conducted using Stata version 18.0. Survey weights and jackknife variance estimation were applied throughout the analysis to account for the complex survey design, ensuring representativeness and providing reliable estimates and standard errors. Descriptive statistics were used to summarize the characteristics of the study population. Continuous variables were presented as means with standard deviations (SD), while categorical variables were reported as frequencies and percentages. Bivariate analysis was performed to explore the independent associations between each independent variable and the outcome variable using the chi-square test of independence (χ²), a p-value of less than 0.10 was considered significant at this level. All variables found to be statistically significant in the bivariate analysis were included in the multivariate analysis. Using bivariable and multivariable logistic regression models, two models were fitted in this study. Model I was an unadjusted model, examining the independent association between the outcome variable and the main exposure variable. Additionally, in Model I each covariate was examined individually to explore its independent association with the outcome variable. Model II was the adjusted model, simultaneously controlling for the main exposure variable and all the covariates. Results in the final model were reported as adjusted odds ratios (aORs) with their 95% confidence intervals (CIs) and statistical significance was set at *p*-value less than 0.05.

Multicollinearity among the independent variables was assessed using the variance inflation factor (VIF), with all explanatory variables showing no significant collinearity (Mean VIF = 1.08, Maximum VIF = 1.19, and Minimum VIF = 1.02). Model fitness was assessed using the Hosmer–Lemeshow goodness of fit test for logistic regression (*p*-value=0.978) demonstrating that the model was well fitted. Findings from this study were reported following the Strengthening Reporting of Observational studies in Epidemiology (STROBE) reporting guidelines.

## Results

### Study participants characteristics

A total of 1,392 unmarried AYA aged 15–24 years who were sexually active were included in the analysis. A majority of the participants were aged 20–24 years (67.9%; *n* = 945) and the mean age was 20.6 ± 2.3 years. Most of the participants were female 51.1% (*n* = 711), while 77.0% (*n* = 1,072) had completed secondary or high school education. Approximately 75% (*n* = 1,038) of the study participants were not employed, while a majority resided in rural areas (79.6%; *n* = 1,108). About 35% (*n* = 490) were from the Manzini region, while more than half had not communicated with parent/guardian on SRH issues (53.9%; *n* = 750). Most of the participants reported to be using contraceptives (75.5%; *n* = 1,051) and 92.5% (*n* = 1,287) reported ever been tested for HIV. At least half of the participants were unaware of PrEP (51.3%; *n* = 714), and a majority reported high confidence in negotiating condom use with their sexual partners (74.7%; *n* = 1,040) (see Table 1).

**Table 1:** Background characteristics of unmarried AYA in Eswatini (Weighted sample N = 1,392)

| Variables | Frequency | Percentage (%) |
| --- | --- | --- |
| *Main exposure variable* |  |  |
| **Alcohol use** |  |  |
| Non-users | 957 | 63.8 |
| Users | 435 | 31.3 |
| *Covariates* |  |  |
| **Age (in years)** (mean = 20.6 years, SD = 2.3) |  |  |
| 15 - 19 | 447 | 32.1 |
| 20 - 24 | 945 | 67.9 |
| **Sex** |  |  |
| Male | 681 | 48.9 |
| Female | 711 | 51.1 |
| **Educational level** |  |  |
| Primary/less | 154 | 11.1 |
| Secondary/Higher | 1,072 | 77.0 |
| Tertiary | 166 | 11.9 |
| **Employment status** |  |  |
| Not employed | 1,038 | 74.6 |
| Employed | 354 | 25.4 |
| **Place of residence** |  |  |
| Rural | 1,108 | 79.6 |
| Urban | 284 | 20.4 |
| **Region** |  |  |
| Hhohho | 330 | 23.7 |
| Lubombo | 275 | 19.8 |
| Manzini | 490 | 35.2 |
| Shiselweni | 297 | 21.3 |
| **Communicated with parent/guardian on SRH matters** |  |  |
| No | 750 | 53.9 |
| Yes | 642 | 46.1 |
| **Contraceptive use** |  |  |
| No | 341 | 24.5 |
| Yes | 1,051 | 75.5 |
| **Ever tested for HIV** |  |  |
| No | 105 | 7.5 |
| Yes | 1,287 | 92.5 |
| **PrEP awareness** |  |  |
| Unaware | 714 | 51.3 |
| Aware | 678 | 48.7 |
| **Condom negotiation confidence** |  |  |
| No confidence | 65 | 4.7 |
| Slight confidence | 73 | 5.2 |
| Moderate Confidence | 214 | 15.4 |
| High Confidence | 1,040 | 74.7 |

### Distribution of Risky Sexual Behaviour

Table 2 shows the distribution of risky sexual behaviour among unmarried AYA by alcohol use and the different covariates. Among alcohol users, a substantial proportion (39.4%; *n* = 289) reported engaging in risky sexual behaviour. Across other variables, the highest proportions of risky sexual behaviour were observed among AYA aged 20–24 years (70.4%; *n* = 517), males (53.4%; *n* = 392), those with secondary or high school education (76.0 %; *n* = 558), unemployed AYA (72.3 %; *n* = 531), AYA residing in rural areas (79.6 %; *n* = 584), AYA from the Manzini region (35.1%; *n* = 258), those who had not discussed SRH matters with a parent/guardian (58.4%; *n* = 429), contraceptives users (71.7%; *n* = 526), AYA who had ever tested for HIV (91.0%; *n* = 668), those aware of PrEP (51.1%; *n* = 375), and AYA having a higher condom negotiation confidence (70.2%; *n* = 515). All variables except place of residence qualified for inclusion in the multivariate analysis (*p* < 0.10).

**Table 2:**
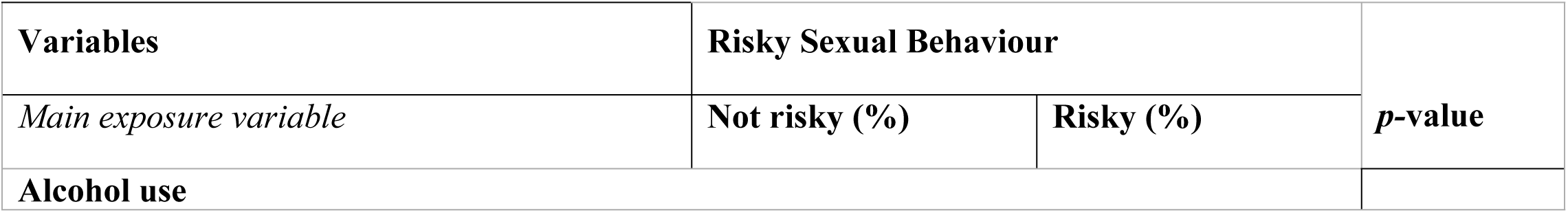
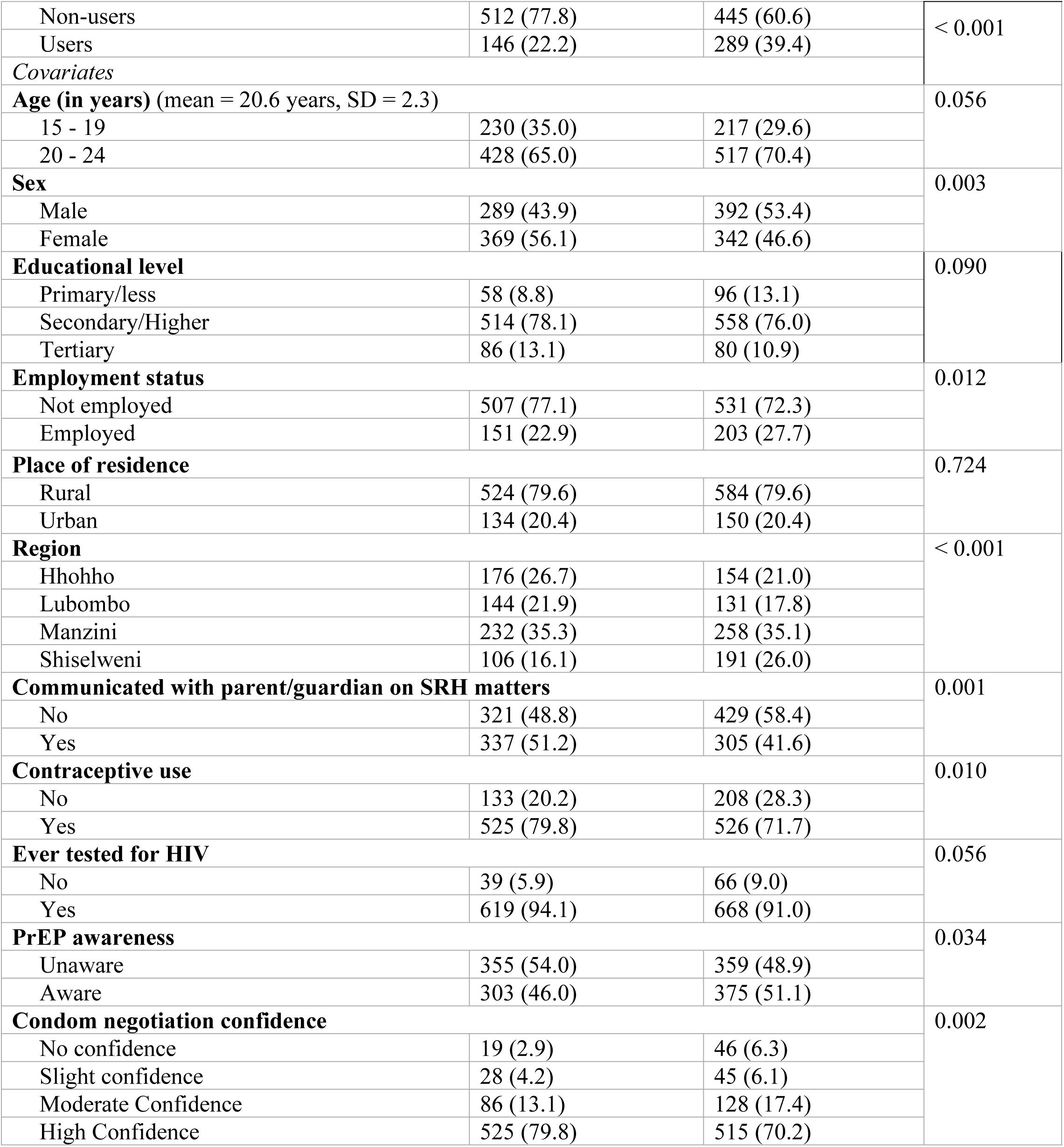
Distribution of risky sexual behaviour by alcohol use and selected covariates among unmarried AYA in Eswatini (Weighted sample N = 1,392)

### The magnitude of risky sexual behaviour and alcohol use among unmarried AYA

The prevalence of risky sexual behaviour among unmarried AYA in this study was 52.7% (95% CI: 50.1 – 55.3) (Fig. 1), while the prevalence of alcohol use was 31.3% (95% CI: 28.9 – 33.7) (Table 1).

**Fig. 1:**
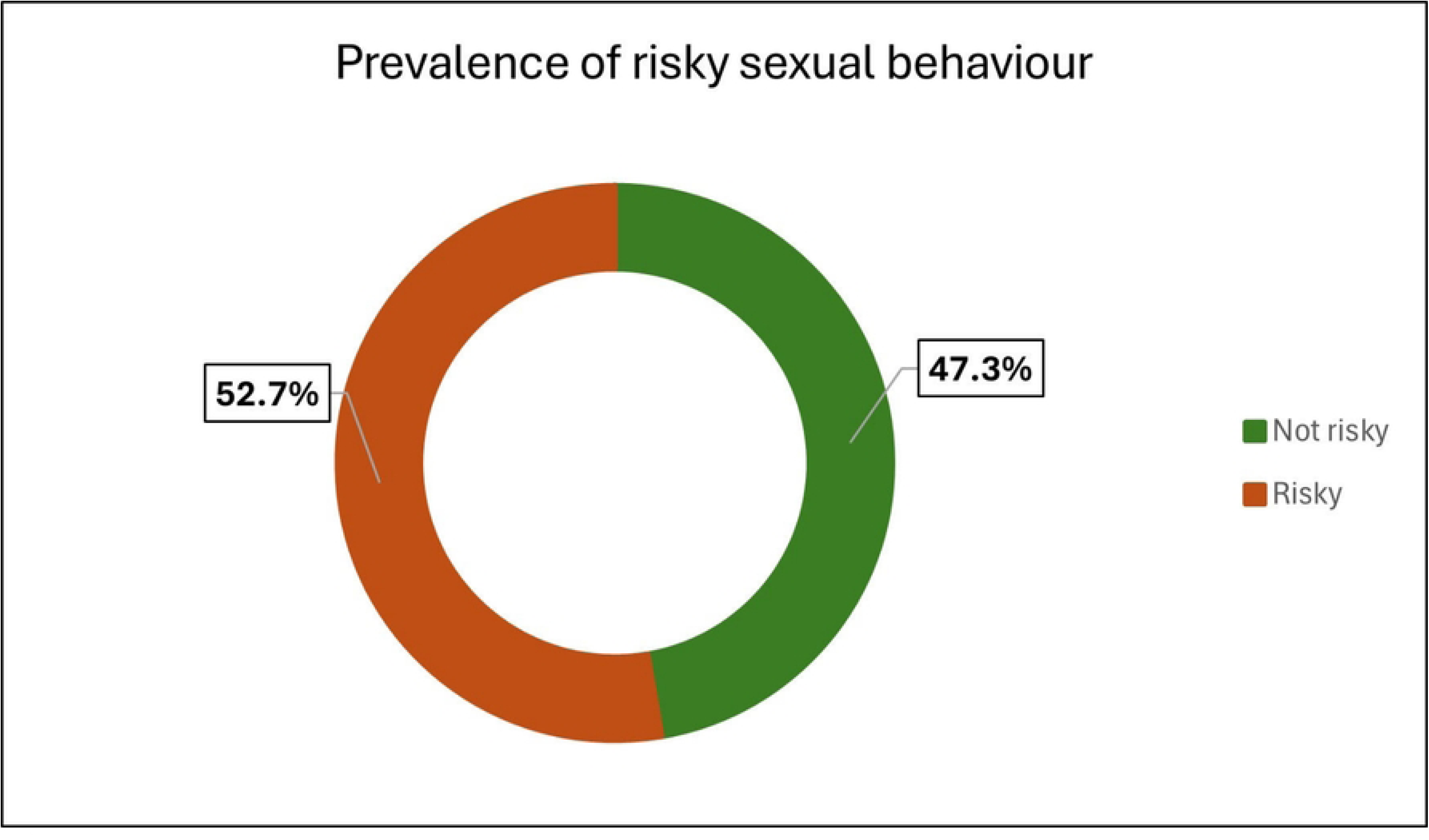
The prevalence of risky sexual behaviour among unmarried AYA in Eswatini

### The association between risky sexual behaviour and alcohol use

The study assessed the association between risky sexual behaviour and alcohol use among unmarried AYA. The results from the unadjusted model (Model I) show that AYA who consume alcohol had significantly higher odds of risky sexual behaviour compared to those who do not consume alcohol (aOR = 2.32, 95% CI: 1.76 – 3.05). In the adjusted model (Model II), the association between risky sexual behaviour and alcohol use remained significant after adjusting for the covariates. The odds of risky sexual behaviour among AYA who consume alcohol were 2.35 (aOR = 2.35, 95% CI: 1.78 – 3.11) times higher compared those who do not consume alcohol, controlling for the other variables in the model (Table 3).

**Table 3:** Determinants of risky sexual behaviour among unmarried AYA Eswatini (Weighted sample N = 1,392)

| Variables | Model I (Unadjusted) | p-value | Model II (Adjusted) | p-value |
| --- | --- | --- | --- | --- |
|  | cOR (95%CI) |  | aOR (95%CI) |  |
| Main Exposure variable |  |  |  |  |
| Alcohol use |  |  |  |  |
| Non-users | Ref |  | Ref |  |
| Users | 2.32 (1.76 – 3.05) | < 0.001 | 2.35 (1.78 – 3.11) | < 0.001 |

| *Covariates* |  |  |  |  |
| --- | --- | --- | --- | --- |
| **Age (in years)** |  |  |  |  |
| 15 - 19 | Ref |  | Ref |  |
| 20 - 24 | 1.26 (1.00 – 1.61) | 0.057 | 1.31 (1.01 – 1.72) | **0.046** |
| **Sex** |  |  |  |  |
| Male | Ref |  | Ref |  |
| Female | 0.69 (0.55 – 0.87) | **0.003** | 0.79 (0.60 – 1.04) | 0.093 |
| **Educational level** |  |  |  |  |
| Primary/less | Ref |  | Ref |  |
| Secondary/Higher | 0.67 (0.47 – 0.97) | **0.034** | 0.66 (0.45 – 0.98) | **0.040** |
| Tertiary | 0.60 (0.38 – 0.93) | **0.026** | 0.56 (0.34 – 0.95) | **0.031** |
| **Employment status** |  |  |  |  |
| Not employed | Ref |  | Ref |  |
| Employed | 1.34 (1.07 – 1.68) | **0.013** | 1.19 (0.93 – 1.53) | 0.165 |
| **Region** |  |  |  |  |
| Hhohho | Ref |  | Ref |  |
| Lubombo | 1.02 (0.69 – 1.50) | 0.937 | 1.04 (0.69 – 1.55) | 0.852 |
| Manzini | 1.34 (1.00 – 1.77) | **0.049** | 1.32 (0.99 – 1.77) | 0.058 |
| Shiselweni | 2.10 (1.53 – 2.89) | **< 0.001** | 2.11 (1.55 – 2.86) | **< 0.001** |
| **Communicated with parent/guardian on SRH matters** |  |  |  |  |
| No | Ref |  | Ref |  |
| Yes | 0.68 (0.55 – 0.85) | **0.001** | 0.72 (0.57 – 0.92) | **0.011** |
| **Contraceptive use** |  |  |  |  |
| No | Ref |  | Ref |  |
| Yes | 0.67 (0.50 – 0.90) | **0.011** | 0.62 (0.45 – 0.86) | **0.005** |
| **Ever tested for HIV** |  |  |  |  |
| No | Ref |  | Ref |  |
| Yes | 0.70 (0.48 – 1.01) | 0.059 | 0.72 (0.46 – 1.12) | 0.142 |
| **PrEP awareness** |  |  |  |  |
| Unaware | Ref |  | Ref |  |
| Aware | 1.28 (1.02 – 1.60) | **0.035** | 1.60 (1.26 – 2.04) | **< 0.001** |
| **Condom negotiation confidence** |  |  |  |  |
| No confidence | Ref |  | Ref |  |
| Slight confidence | 0.60 (0.26 – 1.42) | 0.234 | 0.74 (0.34 – 1.62) | 0.437 |
| Moderate Confidence | 0.56 (0.28 – 1.11) | 0.093 | 0.61 (0.31 – 1.20) | 0.142 |
| High Confidence | 0.37 (0.20 – 0.66) | **0.003** | 0.44 (0.24 – 0.80) | **0.009** |

Ref: Reference category
Bold values indicate statistical significance ( $p < 0.05$ )

### Determinants of risky sexual behaviour among unmarried AYA

After adjusting for covariates, the multivariable logistic regression model (Model II) revealed that the odds of engaging in risky sexual behaviour were higher among AYA aged 20–24 years compared to those aged 15–19 years (aOR = 1.31, 95% CI: 1.01 – 1.72), and that AYA residing in the Shiselweni region had 2.11 (aOR = 2.11, 95% CI: 1.55 – 2.86) times higher odds of engaging in risky sexual behaviour compared to those in the Hhohho region. AYA who were aware of PrEP were 1.60 (aOR = 1.60, 95% CI: 1.26 – 2.04) times more likely to engage in risky sexual behaviour compared to those who were unaware of PrEP. Conversely, the odds of engaging in risky sexual behaviour among AYA who had secondary/high school and tertiary education were 0.66 (aOR = 0.66, 95% CI: 0.45 – 0.98), and 0.56 (aOR = 0.56, 95% CI: 0.34 – 0.95) times lower as compared to AYA with primary or less education, respectively. Compared to AYA who did not have any SRH conversation with a parent or guardian, AYA who communicated with a parent or guardian on SRH matters (aOR = 0.72, 95% CI: 0.57 – 0.92) also had lower odds of engaging in risky sexual behaviour. AYA who used contraceptives had 0.62 (aOR = 0.62, 95% CI: 0.45 – 0.86) times lower odds of engaging in risky sexual behaviour compared to those who did not use contraceptives. Additionally, the odds of engaging in risky sexual behaviour among AYA who had high confidence in negotiating condom use with their sexual partners were 0.44 (aOR = 0.44, 95% CI: 0.24 – 0.80) times lower compared to those who had no confidence (Table 3).

## Discussion

The World Health Organization emphasizes the importance of ensuring that AYA have access to high-quality, adolescent-responsive SRH services, including comprehensive sexuality education, as evidence shows that such can delay the onset of sexual activity, reduce early pregnancies, and improve the reproductive health of AYA [30]. Achieving these goals requires the provision of youth-friendly SRH services, access to contraceptive options for AYA, and reduced societal stigma, especially among unmarried AYA. Therefore, this study sought to determine the prevalence of risky sexual behaviour and its association with alcohol use and identify other determinants of risky sexual behaviour among unmarried AYA Eswatini.

The prevalence of risky sexual behaviour among sexually active unmarried AYA in Eswatini was 52.7% (95% CI: 50.1 – 55.3). This prevalence was higher than that of studies conducted in Rwanda (40%) [31], and Ethiopia (19.5%) [32]. However, the prevalence was lower than that observed in Nigeria (65.8%) [33]. These regional disparities may be due to differences in cultural norms regarding sex, differences in age of sexual debut, exposure to sexual content through media, and access to youth-friendly SRH services across the SSA region. For instance, studies conducted in South Africa and Ghana found that young people without access to a mobile phone or social media were less likely to engage in risky sexual behaviours [34,35], possibly because limited exposure to sexual content reduced the likelihood of sexual experimentation that could occur through risky sexual behaviour.

This study found a significant association between risky sexual behaviour and alcohol use among unmarried AYA. Those who reported consuming alcohol had significantly higher odds of engaging in risky sexual behaviour compared to those who do not consume alcohol (aOR = 2.32, 95% CI: 1.76 – 3.05) in the unadjusted model. The association remained significant even after adjusting for other factors including age, sex, educational status, place of residence, region, communicating with parent/guardian on SRH matters, contraceptive use, HIV testing, PrEP awareness, and confidence in negotiating condom use. In the adjusted model, AYA who consume alcohol were 2.35 (aOR = 2.35, 95% CI: 1.78 – 3.11) times more likely to engage in risky sexual behaviour compared to those who do not consume alcohol. This finding is consistent with studies from other settings [36,37], which also found a higher likelihood of risky sexual practices among alcohol users. This may be explained by the fact that alcohol impairs cognitive judgment and lowers inhibitions, making individuals more likely to engage in unprotected sex, have multiple partners, and fail to negotiate condom use effectively [18,19]. In the context of SSA, alcohol use among young people is often linked to social gatherings such as parties, nightlife, and peer influence, situations that often present increased opportunities for risky sexual behaviour [38,39]. Moreover, alcohol use may serve as a coping mechanism for underlying psychosocial issues that affect young people in low resource settings. The normalization of alcohol consumption in youth culture, combined with limited SRH knowledge and poor access to contraceptive services, creates a high-risk environment for HIV transmission and other negative SRH outcomes [40]. The association between alcohol consumption and risky sexual behaviour underscores the need for integrated interventions that address both substance use and SRH for young people.

The study further revealed that AYA aged 20–24 years were more likely to engage in risky sexual behaviours compared to those aged 15–19 years. This finding aligns with evidence from other studies conducted across sub-Saharan Africa [41–43]. This association may be attributed to the greater autonomy and sexual independence that usually comes with the transition from adolescence to young adulthood. As individuals age, they are more likely to initiate sexual activity, have multiple partners, and engage in relationships with reduced parental supervision and guidance. Additionally, older youth may have increased exposure to environments that facilitate risky sexual behaviours, such as tertiary institutions, nightlife settings, and social gatherings involving alcohol consumption [44–45]. In contrast, younger adolescents are often more closely monitored by parents or guardians and may have less exposure to such high-risk contexts [46]. Tailored age-specific SRH interventions are required, that not only target younger adolescents for early prevention but also address the evolving behavioural and social dynamics influence on sexual risk-taking among older youth and integrate them with life skills, communication, and risk reduction strategies in mitigating sexual risk behaviours among AYA transitioning into adulthood.

The finding that AYA who attained secondary/high school and tertiary education had lower odds of engaging in risky sexual behaviour suggest that higher education equips young people with critical knowledge, decision-making skills, and access to resources that promote safer sexual practices [47]. In low-resource settings, where SRH services may be limited, education serves as an important protective factor by fostering awareness, delaying sexual debut, and empowering youth, especially young women to make informed reproductive health choices [48]. Addressing educational disparities by integrating comprehensive sexuality education into school curricula, while also prioritizing targeted interventions for less-educated youth who remain at elevated risk is necessary. Studies show that higher education levels often correlate with better understanding of SRH, including HIV prevention, contraception, and the consequences of risky sexual practices [49,50]. Therefore, those with higher education are more likely to access SRH services, including counselling, testing, and contraceptives, which reduces risky sexual behaviour.

Living in the Shiselweni region was associated with higher odds of engaging in risky sexual behaviour among AYA compared to residing in the Hhohho region. This may reflect underlying regional disparities in socio-economic conditions, education levels, and access to youth-friendly SRH services. Shiselweni, being more rural, may have fewer recreational facilities and limited structured youth activities, potentially increasing vulnerability to early sexual debut and unprotected sex [51].

Additionally, rural areas often suffer from inadequate healthcare infrastructure, including a shortage of youth-friendly SRH services, and contraceptive options. Cultural norms and stigma surrounding SRH discussions may also be more observed in rural settings, discouraging open communication, further limiting access to accurate information. In contrast, Hhohho, with its more urbanized areas, may offer better educational opportunities, stronger health systems, and more comprehensive youth outreach initiatives, contributing to lower rates of risky sexual behaviour. These regional disparities underscore the urgent need for context-specific interventions in the Shiselweni region that address structural barriers, expand access to SRH services, and create a safe space for youth development.

The finding that having SRH conversations with a parent or guardian is associated with reduced odds of engaging risky sexual behaviours in this study aligns with findings from other SSA countries such as Ethiopia [52,53], and Rwanda [31]. This may be explained that AYA often perceive parents and guardians as trusted sources of accurate information, countering the misinformation that young people may receive from peers and social media. These conversations also foster emotional support and trust, which can empower adolescents to make informed decisions and seek help when needed [54]. In many low-resource settings, particularly in sub-Saharan Africa, where formal SRH education may be limited and cultural taboos often inhibit open discussions about sexuality, family-based communication becomes even more critical. Moreover, discussing SRH within the family context helps establish values that guide behaviour, while also enhancing young people’s ability to negotiate safer sex practices, and resisting peer pressure [55]. The findings underscore the importance of incorporating parent–adolescent communication strategies into SRH programs and policies.

Strengthening parent–adolescent communication should be a central pillar of youth-focused SRH interventions, especially in low-resource and culturally conservative settings.

Consistent with other SSA studies [56], the finding that AYA who reported using contraceptives had significantly lower odds of engaging in risky sexual behaviour suggest that AYA who actively use contraceptives are also more likely to be informed about SRH, practice safer sex, and engage in informed decision-making. Contraceptive use is mostly influenced by awareness of SRH negative outcomes such as HIV, STIs, and unintended pregnancies, this awareness may contribute to the reduced risk of practicing unsafe sexual practices. Moreover, contraceptive users may be more likely to have received SRH education and counselling, which reinforces responsible sexual behaviour. However, this finding is in contrast with a study conducted in Rwanda which found that individuals exposed in risky sexual behaviours do not use contraceptives [31]. The conflicting findings highlight the importance of expanding youth-friendly contraceptive services, integrating them into comprehensive SRH programs, and addressing cultural and systemic barriers that prevent young people from accessing and using contraceptives.

AYA who were aware PrEP had higher odds of engaging in RSB compared to those who were unaware. This finding is contradictory with a study conducted in Côte d’Ivoire which found that PrEP awareness was associated with lower likelihood of engaging in risky sexual behaviours [57].

The differences in these findings may attributed to variations in study period, study population and setting. While PrEP awareness is generally considered as a positive indicator of HIV prevention literacy [58], the observed association in this study may reflect risk compensation which occurs when individuals, perceiving themselves to be protected by biomedical interventions such as PrEP, become less vigilant in practicing other preventive behaviours, such as consistent condom use and limiting the number of sexual partners [59]. This behavioural shift can increase exposure to other sexually transmitted infections and unintended pregnancies, despite reduced HIV risk. These insights underscore the importance of comprehensive risk-reduction counselling when promoting PrEP among youth. Awareness alone is insufficient, it must be paired with robust education on the continued importance of condom use, STI prevention, and healthy relationship dynamics.

Similar to a study conducted in Mozambique [29], the finding that AYA with high confidence in negotiating condom use have lower odds of engaging in risky sexual behaviour suggests that AYA who feel empowered to voice out their preferences and boundaries in sexual relationships are more likely to adopt protective behaviours, such as consistent condom use and avoiding SRH negative outcomes. Confidence in condom negotiation often reflects strong communication skills and personal prioritization, which can be undermined by factors like peer pressure, unequal power dynamics, and fear of rejection [60,61]. Therefore, sexual health interventions targeting youth should go beyond information provision and actively cultivate interpersonal skills, self-esteem, and empowerment. Incorporating role-playing, peer-led discussions, and culturally sensitive approaches can help strengthen negotiation confidence, particularly in settings with high HIV prevalence.

## Conclusion and recommendations

This study demonstrates a significant association between risky sexual behaviour and alcohol use among unmarried AYA in Eswatini. The findings suggest that alcohol consumption substantially increases the likelihood of engaging in unsafe sexual practices, highlighting a critical intersection between substance use and sexual health. Given the potential implications for HIV transmission and other adverse SRH outcomes, these results underscore the urgent need for integrated interventions that address both alcohol use and sexual risk among young people in Eswatini. Tailored interventions that combine substance use prevention, comprehensive sexuality education, and youth-friendly services are essential to promote safer behaviours and improve overall health outcomes for AYA in Eswatini.

## Strengths and limitations

This study offers a number of strengths, including the use of nationally representative data from the Swaziland HIV Incidence Measurement Survey 3 (SHIMS 3), which enhances the generalizability of the findings to unmarried adolescents and young adults in Eswatini. The use of weighted logistic regression models allowed for accurate estimation of associations while controlling for potential confounders. Furthermore, by focusing on the intersection of alcohol use and risky sexual behaviour, the study addresses a critical public health issue and provides actionable insights for youth-focused interventions. However, the study is not without limitations. Its cross-sectional design restricts the ability to establish causality between alcohol use and risky sexual behaviour. Lastly, reliance on self-reported data may be subject to social desirability and recall biases. The dataset also did not look into important psychosocial variables, such as peer influence that could affect sexual behavioural patterns.

## Data Availability Statement

The datasets used in this study are publicly available and can be accessed through the PHIA website at: https://phia-data.icap.columbia.edu/datasets.

## Author Contributions

SGS conceptualized the study, led the data analysis, and drafted the manuscript. AWK, MCS, and COA provided technical guidance on study design and reviewed the manuscript. TPS contributed to data interpretation, drafting and formatting the manuscript, and conducted the literature review. All authors (SGS, AWK, MCS, COA, TPS) reviewed and approved the final version of the manuscript.

## Acknowledgments

We sincerely thank Baylor College of Medicine Childrens Foundation-Mbabane, Eswatini, Siyakhula program initiative for supporting this research. We are also grateful to the African Union Commission (AU), Addis Ababa, Ethiopia through the Pan African University Life and Earth Sciences Institute (including Health and Agriculture), Ibadan, Nigeria for their support in this research. Additionally, we thank the PHIA surveys for permitting us to use the SHIMS 3 dataset, and the adolescents and young adults who participated in the SHIMS 3 survey.

